# Emotion regulation is robustly associated with depressive symptoms across the peripartum – data from a prospective Swedish national cohort study

**DOI:** 10.1101/2024.11.13.24317233

**Authors:** Franziska Weinmar, Emma Fransson, Birgit Derntl, Alkistis Skalkidou

## Abstract

**Importance:** Peripartum depression (PeriPD) is a prevalent condition with serious, long-term consequences for mother and child. Emotion regulation (ER) is increasingly recognized as key factor for maternal mental health and parenting. However, evidence remains limited regarding the relation of ER difficulties and depressive symptoms across the peripartum.

**Objective:** To determine whether self-reported ER difficulties in the antepartum are associated with depressive symptoms at multiple peripartum timepoints. *Secondary*: Whether PeriPD trajectories differ on antepartum ER difficulties.

**Design, Setting and Participants:** Data for this study were collected from January 2022 to April 2024 through the Mom2B cohort, a population-based, prospective Swedish national study using a smartphone app for data collection. The cohort includes Swedish-speaking, pregnant women over 18 years, residing in Sweden, who downloaded the Mom2B app. Eligibility for this sub-study required verification of pregnancy and delivery, along with complete background information (N = 1414). Participants were included if they completed the Difficulties in Emotion Regulation Scale-16 (DERS-16) during the second trimester (N = 623).

**Exposure:** DERS-16 total score at 16-25 weeks antepartum.

**Outcomes and Measures:** Depressive symptoms were assessed using the Edinburgh Postnatal Depression Scale (EPDS) at seven timepoints: 24-34, 36-42 weeks antepartum, and 1-4, 6-13, 14-23, 24-35, 36-42 weeks postpartum. Multiple linear regression models examined DERS-16 scores as exposure, with EPDS scores as outcomes, adjusting for potential confounders. *Secondary*: Comparison of DERS-16 scores between PeriPD trajectories using ANOVA.

**Results:** 623 pregnant women, aged 19-44, were included. DERS-16 scores were strongly associated with EPDS scores up to 14-23 weeks postpartum, even after adjusting for potential confounders (all regression coefficients = .06 - .23, all 95% CIs = .00 - .26, all *p*-values < .05). *Secondary*: 134 participants were classified into a specific PeriPD trajectory. DERS-16 scores differed between trajectories (*F*(4,129) = 26.68, *p* < .001, ηp^2^ = .45), with higher ER difficulties in early and late postpartum-onset trajectories compared to the healthy group (*p* = .017 and *p* = .018, respectively).

**Conclusion and Relevance:** ER difficulties in the second trimester present a robust vulnerability marker for PeriPD symptoms, particularly for progression to postpartum depression. The DERS-16 may aid in early detection of peripartum mental health risks, while enhancing ER offers a promising intervention approach. Further research is needed to evaluate DERS-16’s clinical utility and optimize ER-centered interventions for at-risk trajectories.

**Keypoints:**

- **Question**. Are self-reported difficulties in emotion regulation (ER) in the second trimester of pregnancy associated with depressive symptoms at multiple peripartum timepoints?
- **Findings**. In this population-based prospective Swedish national cohort study, difficulties in ER assessed in the second trimester were significantly associated with higher depressive symptoms during pregnancy and up to six months postpartum, even after adjusting for potential confounders.
- **Meaning**. The study’s findings suggest that ER difficulties during pregnancy present a robust vulnerability marker for depressive symptoms across the peripartum, especially postpartum, and may provide an opportunity for early detection and intervention approaches of peripartum depression.

## Introduction

Peripartum depression (PeriPD), including depressive episodes during pregnancy (antepartum depression, AntePD) or postpartum (postpartum depression, PostPD), is a prevalent mental health condition with serious, long-term consequences for both mother and child.^1-4^ Despite affecting 7-17% of pregnant women and 10-20% of women after childbirth, up to four out of five affected women remain undiagnosed and untreated.^5,6^ Research has identified distinct PeriPD trajectories, such as antepartum-only depression resolving after childbirth, depression with onset in the early or late postpartum period, or persistent symptoms across the peripartum.^7-9^ Although current clinical diagnostic criteria do not yet differentiate PeriPD trajectories^10^, they have been linked to different risk factors and may reflect distinct underlying causes, suggesting the need for tailored diagnostics and interventions.^9,11^

Emotion regulation (ER), a transdiagnostic factor in mental health, is the ability to monitor, understand, and modulate emotional experiences beyond merely suppressing emotions.^12-14^ ER is increasingly recognized as crucial for maternal mental health during the peripartum^15-18^, where maladaptive ER is associated with increased chronic stress^19^, sleep disturbances^20^, substance use^21^, and increased rates of depression, anxiety, and self-injurious thoughts.^18,20-24^ Furthermore, ER may impact parental health, caregiving, and child development.^18,22^ In shaping the parent-child relationship, parental ER capacity is especially important for childhood behavioral development.^25^ To assess ER, studies commonly use the Difficulties in Emotion Regulation Scale (DERS^13^), a validated self-report measure for clinical and non-clinical populations.^13,23,26,27^ The DERS assesses key ER facets: emotional awareness, acceptance, impulse control, goal-directed behavior during negative emotions, and access to effective strategies.^13,28^ It shows strong internal consistency, construct validity, and clinical utility, particularly in peripartum samples, where it correlates well with anxiety and depression measures.^26^ The 16-item short form, DERS-16^29^, retains excellent psychometric properties, offering a brief yet reliable alternative for pregnant samples in time-constrained settings.^27,29^

As a modifiable ability, ER is critical for assessing mental health risks and serves as a potential intervention target during the peripartum.^30^ However, research on how ER during pregnancy relates to depressive symptoms and PeriPD trajectories is limited. Addressing this gap, the current study investigated ER using the DERS-16 during the second trimester of pregnancy and their association with depressive symptoms at multiple peripartum timepoints, using data from a population-based, prospective Swedish national cohort study. Additionally, we analyzed ER differences across PeriPD subgroups based on onset and persistence of symptoms. This study ultimately aims to assess the utility of the DERS-16 as a concise screening tool during pregnancy, supporting PeriPD risk assessment and guiding prevention and intervention strategies.^26,28,29,31^ We hypothesized ER difficulties reported in the second trimester to be associated with elevated depressive symptoms across the peripartum^23,24,26,27^, and given that distinct PeriPD trajectories are associated with diverse background factors^9^, we expected ER capacity to vary between PeriPD trajectories.

## Methods

### Participants and procedure

Data were obtained from the Mom2B cohort (www.mom2b.se), an ongoing, prospective Swedish national study using a smartphone-app for data collection.^32^ All Swedish-speaking women over 18 residing in Sweden, who are pregnant or within three months postpartum, can participate by downloading the Mom2B app. Recruitment occurs via healthcare facilities, social media, and print advertisements. After providing informed consent, participants complete online surveys accessible at designated peripartum periods.^32^ Pregnancy and delivery are confirmed via the Swedish national birth registry. The study complies with General Data Protection Regulations and has ethical approval from the Swedish Ethical Review Committee (dnr: 2019/01170, with amendments). Compared to Sweden’s general pregnant population, the Mom2B cohort has a higher proportion of highly educated individuals and a lower proportion of participants born outside Sweden.^32-34^ This study used data collected between 01/2022 to 04/2024, specifically including responses to the DERS-16^29^ which was included from 05/2022 on and was available for participants at 16-25 weeks antepartum. Eligibility criteria included verified pregnancy and delivery, complete background information (N = 1414), and a completed DERS-16 during the specified period (N = 623). The Mom2B app allows users to join at any pregnancy stage, but survey access is limited to specific time windows, which can result in missing data–a common issue in mobile health research^35^

### Sociodemographic information

In the background questionnaire participants reported in the app country of birth, age at registration, highest education level (“no schooling”, “primary school”, “high school”, “polytechnic/vocational training”, “university or college”), mental health history (“no”, “yes, with professional help”, “yes, without professional help”), and parity. Additional information included height and weight (body-mass-index (BMI) calculation), relationships status (“no partner”, “partner, cohabiting”, “with a partner, no cohabiting”), partner violence history, substance use three months before pregnancy (alcohol, cigarettes, snus), mental health issues due to oral contraceptives (OC), past treatment for premenstrual disorders, and pregnancy loss history (all “yes”/”no”). After birth, participants reported mode of delivery (“vaginal delivery”, “assisted vacuum delivery”, “planned caesarean section”, “emergency caesarean section”) and neonatal issues up to 2 weeks postpartum (“yes”/”no”). Breastfeeding was tracked up to 42 weeks postpartum (“yes, full/partly”, “no”).

### Difficulties in Emotion Regulation Scale-16

Emotion regulation was measured with the Difficulties in Emotion Regulation Scale-16 (DERS-16^29^), a 16-item short form of the DERS^13^, at 16-25 weeks antepartum and 17-25 weeks postpartum. The DERS-16 assesses trait-level emotion dysregulation using five subscales: : (1) Lack of emotional *clarity*, e.g., “I have difficulty making sense out of my feelings”; (2) *Non-acceptance* of emotional responses, e.g., “When I’m upset, I become angry at myself fore feeling that way”; (3) *Impulse* control difficulties, e.g., “When I’m upset, I become out of control”; (4) Difficulty in engaging in *goal-directed behavior*, e.g., “When I’m upset, I have difficulty getting work done”; (5) Limited access to emotion regulation *strategies*, e.g., “When I’m upset, I believe there is nothing I can do to feel better”.^27-29^ Scores range from 16 to 80, with higher scores indicating greater emotion dysregulation.^13,29^ The DERS-16 has shown strong psychometric properties, including high internal consistency, test-retest reliability, and convergent and discriminant validity^29^, also in peripartum samples.^26^

### Edinburgh Postnatal Depression Scale

PeriPD symptoms were measured using the Swedish version of the Edinburgh Postnatal Depression Scale (EPDS^36-38^) at eight timepoints: 12-22, 24-34, and 36-42 weeks antepartum as well as 1-4, 6-13, 14-23, 24-35, and 36-42 weeks postpartum. The EPDS is a validated 10-item self-report screening tool assessing depressive symptoms over the past seven days, with higher scores indicating greater severity.^37,38^ Scores of ≥13 antepartum and ≥12 postpartum indicate clinically relevant symptoms, as validated in Swedish samples.^37,38^ These cut-off scores were also used for the present secondary analyses in the PeriPD trajectories^9^. If respondents completed nine of ten items, the missing score was imputed with the mean of the other items; with fewer than nine completed items, the total score was set to missing.

### Additional psychometric surveys

Based on a study using data from a population-based prospective cohort study and machine learning methods to predict depressive symptoms six weeks postpartum^39^ we included additional psychometric scales provided via the Mom2B app during antepartum as potential confounders for our primary analyses. The extra surveys included the Fear of Birth Scale (FOBS^40^), Resilience Scale-14 (RS-14^41,42^), Sense of Coherence Scale-13 (SOC-13^43,44^), and the Vulnerable Personality Style Questionnaire (VPSQ^45,46^). Delivery experience was assessed via a visual analogue scale with low values indicating a negative and high values indicating a positive delivery experience.

### Statistical analyses

For all statistical analyses, IBM SPSS Statistics (version 28.0) was used and the alpha criterion level set to *p* ≤ .05.

### Emotion regulation across the peripartum

To assess emotion regulation across peripartum, we performed a Pearson correlation analysis of DERS-16 total score at 16-25 weeks antepartum with scores at 17-25 weeks postpartum. A paired samples t-test was conducted to test if DERS-16 total scores were different in the postpartum compared to the antepartum assessment period.

### Primary analyses: Association of ER and peripartum depressive symptoms

To assess the association of ER and depressive symptoms, we performed multiple linear regression models with the DERS-16 total score at 16-25 weeks antepartum as the exposure and EPDS total score across peripartum (24-34, 36-42 weeks antepartum; 1-4, 6-23, 24-35, 36-42 weeks postpartum), as outcomes, respectively. Model 1 was unadjusted for any other potential confounders; Model 2 adjusted for potential confounders including age, pre-pregnancy BMI, university-level of education, parity, pregnancy loss history, self-reported history of depression, treatment for premenstrual disorder, mental health issues from OC use, mean FOBS, RS-14, SOC-13, and VPSQ scores (all assessed antepartum). Postpartum EPDS outcomes were additionally adjusted for delivery experience, neonatal issues (0-2 weeks postpartum) and in Model 3 also adjusted for EPDS score at 12-22 weeks antepartum.

### Secondary analyses: ER across PeriPD trajectories

To explore ER differences across PeriPD trajectories, we categorized participants into five groups based on EPDS scores above the clinical cut-offs (≥13 antepartum, ≥12 postpartum) following Wikman et al.^9^: (1) healthy (no depressive symptoms antepartum or postpartum), (2) antepartum-only depression, (3) early postpartum-onset depression, (4) late postpartum-onset depression, and (5) persistent depression. Group differences between EPDS responders and non-responders (those completing DERS-16 but missing any EPDS assessment) were tested using t-tests or Chi-squared tests. Trajectory group differences were analyzed with Fisher’s Exact test for categorical variables, ANOVA, or Kruskal-Wallis H tests, if homogeneity of variance was violated. To assess DERS-16 score differences by PeriPD trajectory, univariate ANOVA was used, with effect sizes reported as ηp^2^ and Bonferroni-adjusted post-hoc tests for significant effects.

## Results

### Sample characteristics

In total, 623 participants from the Mom2B study completed the DERS-16 at 16-25 weeks antepartum and were included in this study (see *Figure 1*). Sample characteristics are provided in *Table 1*. Due to varying availability of surveys across peripartum periods, sample sizes differ for each analysis (see *Table 1*). Among those completing the EPDS, depressive symptoms above the cut-off (score of ≥ 13) were noted in 18.9% at 24-35 weeks and 15.5% at 36-42 weeks antepartum. Postpartum, EPDS scores (≥ 12) were observed as follows: 23.6% at 1-4 weeks, 17.4% at 6-13 weeks, 16.7% at 14-23 weeks, 15.9% at 24-35 weeks, and 14.8% at 36-49 weeks, reflecting clinically relevant depressive symptoms.^37,38^

**Table 1.** Characteristics of study sample.

| Characteristics | N (%)<br>Responders | Mean<br>(SD) | Range<br>Min-Max |
| --- | --- | --- | --- |
| **Total** | **623 (100%)** |  |  |
| Age (years) | 623 (100%) | 31.66<br>(4.57) | 19-44 |
| Pre-pregnancy BMI (kg/m<sup>2</sup>) | 616 (98.9%) | 26.07<br>(6.04) | 14-69 |
| Education | 623 (100%) |  |  |
| University level education | 423 (67.9%) |  |  |
| Relationship status | 623 (100%) |  |  |
| Partner, not cohabiting | 606 (97.3%) |  |  |
| Partner, cohabiting | 585 (93.9%) |  |  |
| Partner violence | 596 (95.7%) |  |  |
| Current partner violence | 11 (1.8%) |  |  |
| Previous partner violence | 140 (23.5%) |  |  |
| Mental health history | 623 (100%) |  |  |
| Depression history (self-reported) | 340 (54.6%) |  |  |
| Professional help | 276 (44.3%) |  |  |
| Anxiety history (self-reported) | 275 (44.2%) |  |  |
| Professional help | 201 (32.3%) |  |  |
| Female-specific mental health | 618 (99.2%) |  |  |
| Treatment for premenstrual disorders | 62 (10%) |  |  |
| Mood swings from oral contraceptives | 308 (49.8%) |  |  |
| Substance use | 617 (99.2%) |  |  |
| Alcohol, 3 months before pregnancy | 478 (77.5%) |  |  |
| Alcohol, ≥ 7 glasses/week<br>3 months before pregnancy | 13 (2.7%) |  |  |
| Smoking, 3 months before pregnancy | 85 (13.8%) |  |  |
| Snus, 3 months before pregnancy | 112 (18.0%) |  |  |
| **Pregnancy-related variables** |  |  |  |
| Pregnancy week at registration to Mom2B study | 603 (96.79%) | 31.66<br>(4.57) | 2-26 |
| Parity | 606 (97.3%) |  |  |
| Primiparous | 234 (38.6%) |  |  |
| History of pregnancy loss | 610 (97.9%) |  |  |
| Previous pregnancy loss | 206 (33.8%) |  |  |
| Fear of Birth Scale<br>(mean total score 13-42 w antepartum) | 608 (97.6%) | 38.31<br>(25.49) | 0-100 |
| **Delivery-related variables** |  |  |  |
| Delivery experience 0-2 w postpartum<br>(scale from 0 = negative to 100 = positive) | 357 (57.3%) | 76.6<br>(20.16) | 0-100 |
| Mode of delivery | 357 (57.3%) |  |  |
| Vaginal delivery | 263 (73.7%) |  |  |
| Assisted vacuum delivery | 29 (8.1%) |  |  |
| Planned caesarean section | 29 (8.1%) |  |  |
| Emergency caesarean section | 36 (10.1%) |  |  |
| **Postpartum-related variables** |  |  |  |
| Neonatal issues | 356 (57.1%) |  |  |
| Reported neonatal issues up to 0-2 w postpartum | 43 (12.1%) |  |  |
| Breastfeeding | 340 (54.6%) |  |  |
| Full/partly | 337 (99.1%) |  |  |
| **Psychometric scales** |  |  |  |
| DERS-16 (total score) 16-25 w antepartum | 623 (100%) | 32.42<br>(13.03) | 16-76 |
| DERS-16 (total score) 17-25 w antepartum | 199 (31.9%) | 32.45<br>(13.67) | 16-73 |
| SOC-13 (total score) 18-42 w antepartum | 565 (90.7%) | 55.61<br>(6.70) | 31-78 |
| RS-14 (total score) 20-42 w antepartum | 550 (88.3%) | 75.41<br>(12.36) | 28-98 |
| VPSQ (total score) 32-42 w antepartum | 420 (67.4%) | 26.95<br>(4.09) | 19-41 |
| EPDS (total score) 12-22 w antepartum | 545 (87.5%) | 7.45<br>(5.19) | 0-27 |
| EPDS (total score) 24-34 w antepartum | 518 (83.1%) | 7.50<br>(5.19) | 0-26 |
| EPDS (total score) 36-42 w antepartum | 373 (59.9%) | 6.82<br>(5.29) | 0-24 |
| EPDS (total score) 1-4 w postpartum | 339 (54.4%) | 7.99<br>(5.47) | 0-26 |
| EPDS (total score) 6-13 w postpartum | 282 (45.23%) | 6.34<br>(5.06) | 0-24 |
| EPDS (total score) 14-23 w postpartum | 221 (35.5%) | 6.38<br>(5.16) | 0-25 |
| EPDS (total score) 24-35 w postpartum | 151 (24.2%) | 5.37<br>(5.09) | 0-21 |
| EPDS (total score) 36-49 w postpartum | 81 (13.0%) | 5.62<br>(5.03) | 0-19 |

w = weeks
*Note.* Results are presented as frequencies and relative frequencies within the subset of survey-responders and if applicable additionally means (standard deviation, SD). BMI: Body mass index; DERS-16: Deficits in Emotion Regulation Scale-16; EPDS: Edinburgh Postnatal Depression Scale; RS-14: Resilience Scale-14; SOC-13: Sense of Coherence-13; VPSQ: Vulnerable Personality Style Questionnaire.

**Figure 1.**
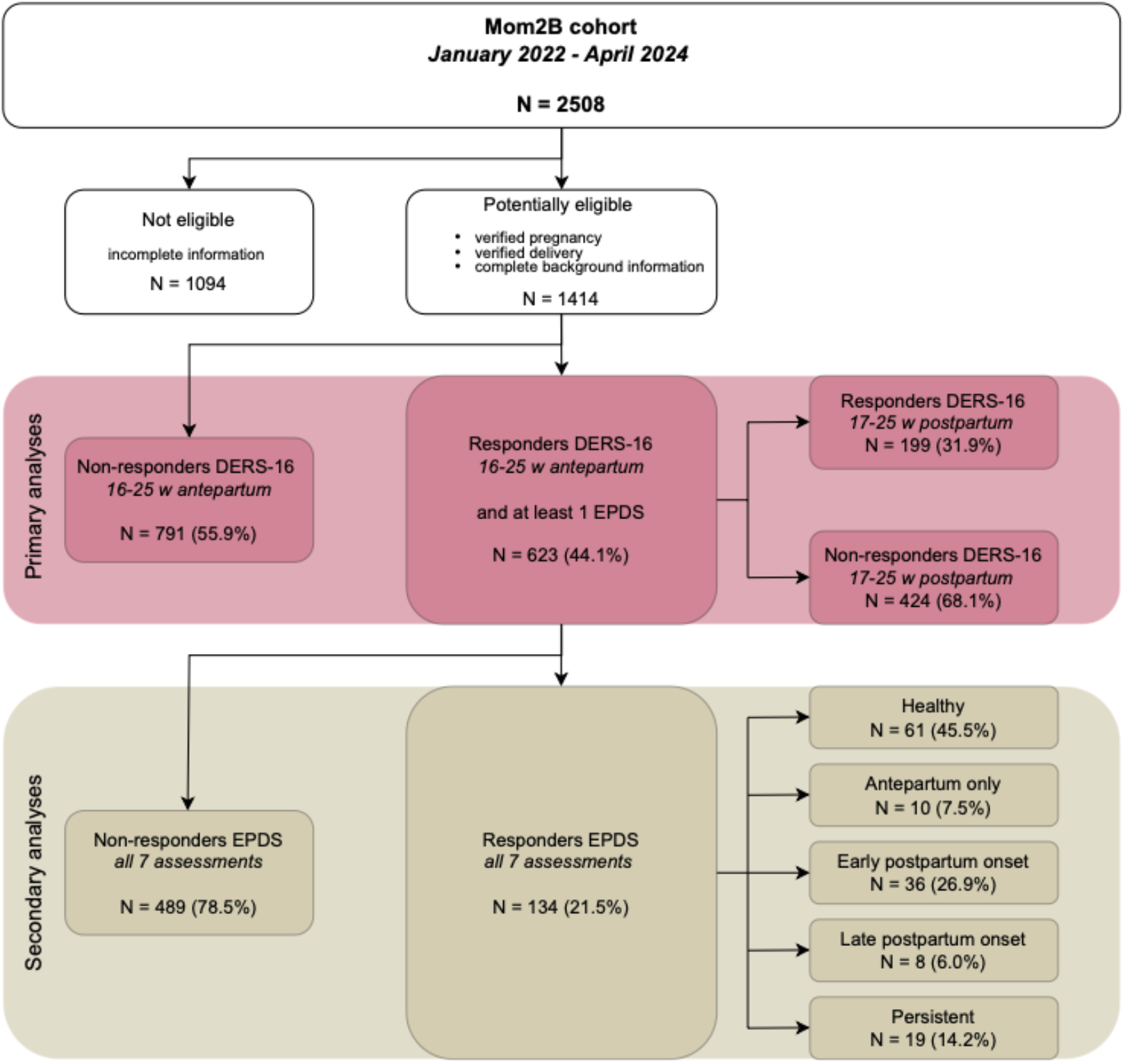
Flowchart of included participants in the study. Note. Data for the present study were derived from January 2022 to April 2024 from the prospective Swedish national cohort study Mom2B (35) as the Difficulties in Emotion Regulation Scale-16 (32) was introduced from May 2022 and was available for participants at 16-25 weeks antepartum.

### Emotion regulation remains stable over the peripartum

For stability analysis, 199 participants filled in the DERS-16 for a second time at 17-25 weeks postpartum (see *Figure 1*). DERS-16 scores at 16-25 weeks antepartum were strongly correlated with scores at 17-25 weeks postpartum (*r*(197) = .655, *p* <.001) with no significant mean differences between periods (*p* = .076, see *Supplementary Information*).

### Emotion regulation is robustly associated with peripartum depressive symptoms

Regression models (see *Table 2*) showed that DERS-16 score at 16-25 weeks antepartum (exposure) is significantly associated with peripartum EPDS scores (outcome). DERS-16 was significantly associated with EPDS scores until 14-23 weeks postpartum in all three regression models (*see Table 2:* crude *p*-values < .001; adjusting for potential confounders excluding EPDS antepartum *p*-values < .017, adjusting for potential confounders including EPDS antepartum *p*-values < .05). While DERS-16 was significantly associated with EPDS assessed at 24-35 weeks postpartum in the crude model, this association became non-significant with the coefficients attenuated towards the null when adjusting for potential confounders (excluding EPDS antepartum: *p* = .111; including EPDS antepartum: *p* = .517). For EPDS scores at 36-49 weeks postpartum, the association was significant in the crude model (*p* < .001) and the confounder-adjusted model (excluding EPDS antepartum: *p* = .018), but non-significant after adjusting for EPDS antepartum (*p* = .196). *Figure 2* shows all included regression coefficients (exposure and potential confounders) and color-coded their significant positive or negative association with EPDS score across the peripartum. All regression coefficients and associated statistics for potential confounders are reported for the different regression models in *Supplementary Table 1*.

**Table 2.** Regression coefficients and 95% confidence interval of Deficits of Emotion Regulation Scale-16 (DERS-16) 16-25 weeks antepartum (exposure) in linear regression models on Edinburgh Postnatal Depression Scale (EPDS) across the peripartum period (outcome).

| (Outcome) | (1) | (2) | (3) |
| --- | --- | --- | --- |
| EPDS total score across the peripartum period | Unadjusted | Adjusted for potential confounders – EPDS total score 12-22 w antepartum | Adjusted for potential confounders + EPDS total score 12-22 w antepartum |
| **beta coefficients for DERS-16 total score 16-25 w antepartum** |  |  |  |
| 24-34 w antepartum | .22 (.19 to .25) *** | .15 (.11 to .19) *** | .15 (.11 to .19) *** |
| 36-42 w antepartum | .23 (.19 to .26) *** | .15 (.11 to .20) *** | .15 (.11 to .20) *** |
| **BIRTH** |  |  |  |
| 1-4 w postpartum | .17 (.13 to .22) *** | .08 (.03 to .13) *** | .06 (.00 to .11) * |
| 6-13 w postpartum | .18 (.13 to .22) *** | .09 (.04 to .14) *** | .06 (.00 to .11) * |
| 14-23 w postpartum | .21 (.17 to .26) *** | .15 (.09 to .20) *** | .10 (.04 to .15) *** |
| 24-35 w postpartum | .15 (.10 to .21) *** | .06 (-.01 to .13) + | .03 (-.05 to .10) *ns* |
| 36-49 w postpartum | .22 (.15 to .29) *** | .09 (.02 to .16) * | .04 (-.02 to .11) *ns* |

*ns* $p \geq .196$ , + $p = .111$ , \* $p < .050$ , \*\*\* $p \leq .001$ ; w: weeks
*Note.* Models are corrected for (1) no other potential confounders (2) potential confounders excluding EPDS total score 12-22 weeks antepartum and (3) potential confounders including EPDS total score 12-22 weeks antepartum. Results for all beta coefficients can be found in *Supplementary Table 1*.

**Figure 2.**
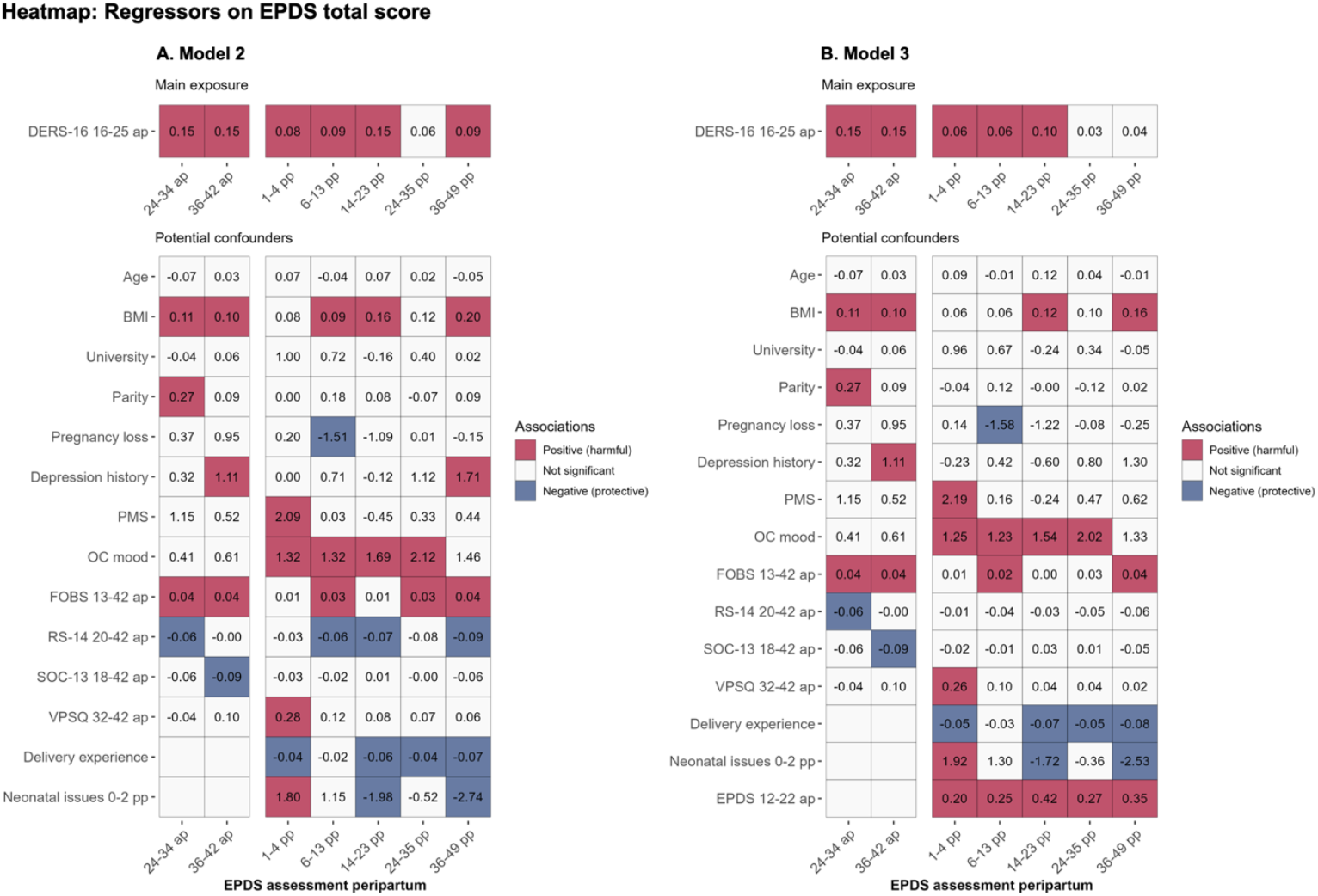
Regression coefficients in linear regression models on Edinburgh Postnatal Depression Scale (EPDS) total score across peripartum (outcome). Note. A. Model 2 with Deficits of Emotion Regulation Scale-16 (DERS-16) 16-25 weeks antepartum (exposure) and other potential confounders excluding EPDS total score 12-22 weeks antepartum. B. Model 3 with DERS-16 16-25 weeks antepartum (exposure) and other potential confounders including EPDS total score 12-22 weeks antepartum. Heatmaps show regressors and their respective coefficient value on EPDS total scores across peripartum in the tiles. The color of the tiles is dependent on the regressors’ significance and direction of association (positive or negative). DERS-16 16-25 ap: Deficits of Emotion Regulation Scale- 16 total score at 16-25 weeks antepartum; BMI: pre-pregnancy body-mass-index; University: university level education (vs less); Pregnancy loss: pregnancy loss history (vs never); Depression history: self-reported depression history with professional help (vs no); PMS: past treatment for premenstrual disorder (vs never); OC mood: mental health issues due to oral contraceptives (vs never); FOBS 13-42 ap: Fear of Birth Scale, mean total score at 13-42 weeks antepartum; RS-14 20-42 ap: Resilience Scale-14 total score at 20-42 weeks antepartum; SOC-13 18-42 ap: Sense of coherence-13 total score 18-42 weeks antepartum; VPSQ 32-42 ap: Vulnerable Personality Style Questionnaire total score at 32-42 weeks antepartum; Delivery experience: delivery experience at 0-2 weeks postpartum; Neonatal issues 0-2 pp: neonatal issues up to two weeks postpartum (vs none). EPDS 12-22 ap: Edinburgh Postnatal Depression Scale total score at 12-22 weeks antepartum.

**Figure 3.**
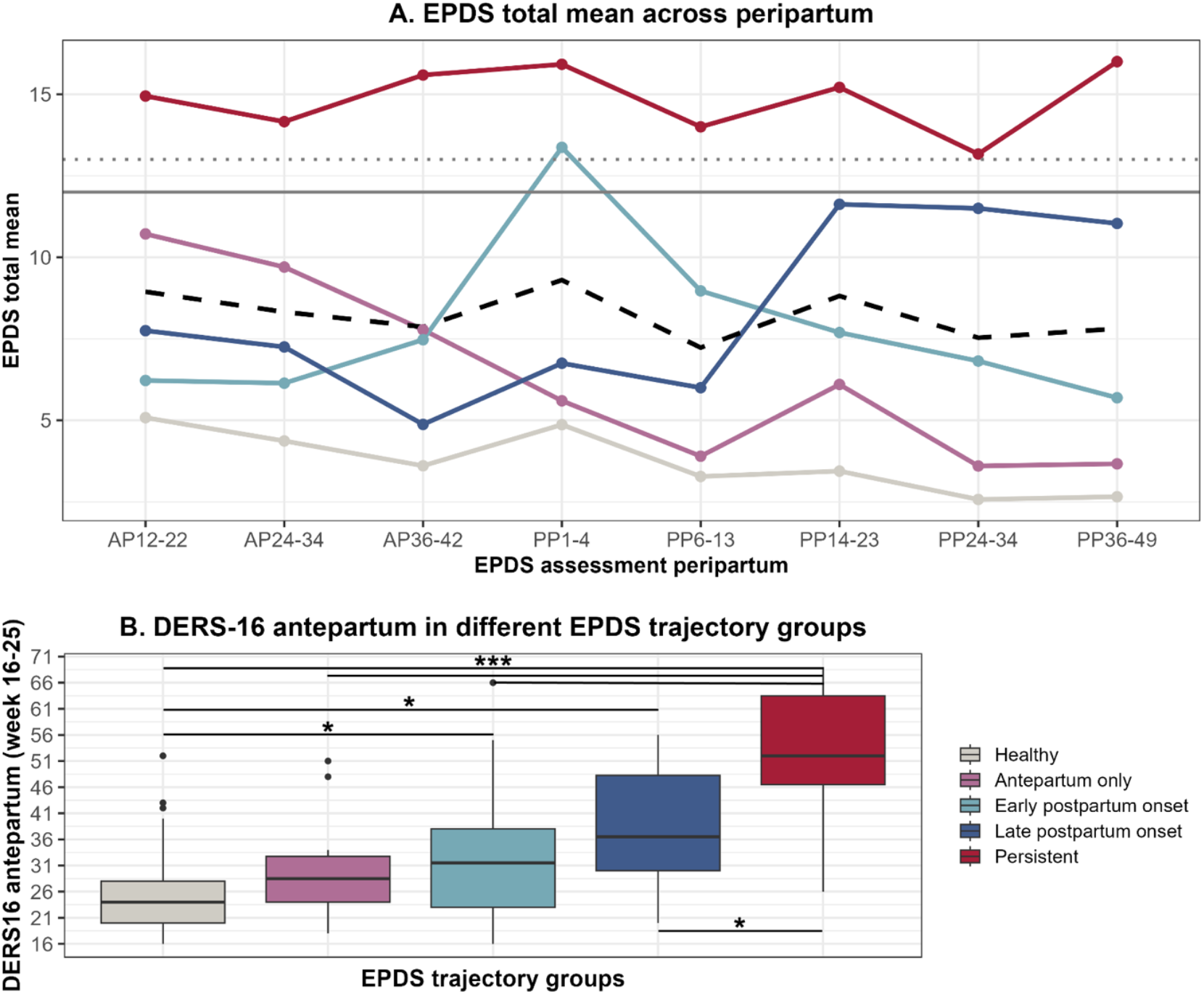
Peripartum depression trajectories and differences in Deficits of Emotion Regulation Scale-16 (DERS-16) total score. Note. A. Edinburgh Postnatal Depression Scale (EPDS) mean total scores for each assessment period across peripartum shown for five distinct peripartum depression trajectory groups. The overall EPDS mean score across the groups is represented by the black dashed line. The grey reference lines indicate the EPDS cut-off score for depression with a score of 13 (dotted) used in antepartum and score of 12 (solid) used in postpartum. B. Differences in in Deficits of Emotion Regulation Scale-16 (DERS-16) mean total scores at 16-25 weeks antepartum (N = 134) between the five distinct peripartum depression trajectory groups. Boxes show upper and lower quartile values with the median indicated as bold line. Whiskers show minimum and maximum scores. *p < .005, ***p < .001

### Emotion regulation differs across PeriPD trajectories

Of the 623 participants, 134 completed the EPDS across all seven timepoints, enabling PeriPD trajectory classification. Compared to the complete EPDS responders, complete EPDS non-responders registered later in pregnancy (*t*(601) = −.2.69, *p* < .001), had a higher pre-pregnancy BMI (*t*(614) = −.55, *p* = .041) and had more planned caesarean sections (X^2^(3) = 8.93, *p* = .030). No other differences were found (all *p-*values ≥ .078). Trajectories referred to: healthy (45.5%), antepartum-only (7.5%), early postpartum-onset (26.9%), late postpartum-onset (6.0%), and persistent depression (14.2%; see *Figure 1*). Significant differences in antepartum DERS-16 scores were found between the PeriPD trajectory groups (*F*(4,129) = 26.68, *p* < .001, ηp^2^ = .45; *see Figure 4*). Post-hoc tests showed higher antepartum DERS-16 scores in early and late postpartum-onset trajectories compared to the healthy group (*p* = .017, *p* = .018, respectively), with the persistent depression trajectory having higher DERS-16 scores than all other groups (all *p*-values ≤ .006). *Supplementary Tables 2* and *3* provide descriptive information.

## Discussion

This Swedish national app-based cohort study showed that self-reported emotion regulation (ER) remained relatively stable from pregnancy through the postpartum period. Difficulties in ER, assessed in the second trimester via the DERS-16, were strongly associated with peripartum depression (PeriPD) symptoms up to 14-23 week postpartum, even after adjusting for potential confounders. This association, however, did not extend robustly beyond six months postpartum when accounting for antepartum depression (AntePD) scores. Notably, women who would later meet the EPDS threshold for postpartum depression (PostPD) already displayed increased ER difficulties as early as the second trimester, pinpointing a possibility for early identification of high-risk individuals.

The DERS-16 scores as reported in pregnancy and postpartum showed a strong correlation without significant mean changes, supporting its reliability for assessing ER across the peripartum period. This aligns with previous findings on the stability of ER^29^ also during the peripartum period.^19,24,47^ While Coo et al.^24^ reported overall stability of ER from pregnancy to postpartum and observed improvements in DERS subscales, our findings indicate a tendency toward slightly higher, i.e. worse, DERS-16 scores four to six months postpartum, though not statistically significant. This discrepancy may relate to the assessment timing differences (Coo et al. third trimester, period of heightened bodily changes and symptoms as well as imminent psychosocial changes; current study second trimester, women may not yet be experiencing elevated stress to this extent) and ER assessment versions (Coo et al. used an adapted version of DERS^48^ and analyzed subscale scores; current study used DERS-16 total score). Despite the overall stability, both studies highlight subtle variability in ER that could be addressed through targeted interventions. Generally, our findings highlight DERS-16’s utility as a reliable tool for assessing ER difficulties across the peripartum.

ER difficulties assessed in the second trimester were strongly associated with depressive symptoms throughout the peripartum and up to six months postpartum, even after controlling for potential confounders, supporting previous evidence linking ER to psychopathology and mental health vulnerability.^12,49^ Our findings are in line with previous studies showing an association between emotion dysregulation and higher levels of AntePD and PostPD symptoms.^16,20,22,23,50,51^ Unlike prior studies that found no significant link between antepartum ER and PostPD^23^, our data show a robust association across multiple postpartum timepoints. This association, however, weakened beyond six months postpartum, suggesting the DERS-16’s sensitivity for depressive risk might be limited after this period. Our results underscore ER as an important factor in identifying PeriPD risk and support the clinical utility of the DERS-16 as a prognostic tool during pregnancy.

Remarkably, the early and late postpartum-onset depression trajectories reported significantly more ER difficulties during the second trimester, even while still below the threshold for depression during the time of the ER assessment. This finding aligns with Wikman et al.^9^, who identified distinct characteristics among PeriPD trajectories before childbirth, yet extending this work by demonstrating that these trajectories may differ in psychological abilities like ER. Recognizing emotion dysregulation as an early vulnerability marker for PostPD is particularly relevant for the late postpartum-onset trajectory, where many cases go undiagnosed, partly due to diagnostic criteria not acknowledging symptom onset beyond six weeks postpartum.^10,52^ Thus, assessing ER difficulties during pregnancy with the 16-item self-report scale could be of considerable value for healthcare providers. Contrary to our expectations, the antepartum-only depression trajectory showed no significant differences in DERS-16 scores compared to the healthy group, suggesting that women in the antepartum-only trajectory may have had adaptive ER skills, limiting symptoms to pregnancy only. Moreover, our data imply that the antepartum-only trajectory might not be primarily linked to emotion dysregulation. However, this lack of difference in self-reported ER warrants replication, partly due to small sample size in the antepartum-only group. In contrast, the persistent trajectory showed significantly higher ER difficulties during pregnancy, with depressive symptoms persisting throughout the peripartum period, indicating sustained vulnerability. Our study highlights the heterogeneity of PeriPD, challenging the traditional dichotomous view of this condition. A nuanced understanding is vital for identifying trajectory or subgroup-specific causes and risk factors.^9^ Ultimately, effectively communicating the distinct mental health risks and needs associated with different PeriPD trajectories, such as ER, to healthcare providers is essential^9^, paving the way for personalized medical approaches to prevention and treatment.

Although this study features a large, population-based and well-characterized prospective sample, some limitations should be noted: First, the cohort’s overrepresentation of Swedish-born women with higher education may limit generalizability of findings, particularly given the higher prevalence of PeriPD among women from lower socioeconomic or minority backgrounds.^53,54^ Second, high rates of self-reported history of mental illness potentially introduces a level of selection bias, as individuals more attuned to mental health concerns may be more likely to participate in a study on maternal mental health. Third, missing data and non-response at different timepoints, a common challenge of mobile health research^35^, led to variable sample size in our regression analyses, which could be linked to how well or poorly participants felt during the peripartum period. Also, dropout rates during later postpartum timepoints increased. These issues, however, were mitigated by maintaining a fairly large sample size and ensuring similar background characteristics between responders and non-responders. The EPDS, although a widely used screening tool, is not a diagnostic instrument and has been discussed to capture general psychological distress rather than specifically just depressive symptoms.^55-57^ Also, we did not evaluate symptom severity in relation to clinically diagnosed depression, nor account for potential treatment effects that might have altered symptom trajectories. Instead, our goal was to examine the association between ER and the EPDS as a measure of mental health and well-being. Our results underscore ER’s role as a transdiagnostic factor for mental health, highlighting it as a tangible target for prevention and intervention efforts. Finally, PeriPD is a multifactorial disorder^8,58,59^, with ER likely representing just one part of its complex etiology. While future research should approach PeriPD from multiple perspectives, our study’s strength lies in addressing this complexity by controlling for diverse known and available confounders in the Mom2B dataset.

In conclusion, this study emphasizes the significance of ER difficulties during the second trimester of pregnancy as an early and robust vulnerability marker for PeriPD symptoms, particularly for cases progressing to PostPD. The findings endorse the DERS-16 as a practical screening tool for identifying peripartum mental health risks and guiding timely interventions.^26,27^ Given that effective ER skills can be trained and strengthened during pregnancy^18,60,61^, enhancing ER during pregnancy presents a promising avenue for prevention and intervention. Future research should evaluate the clinical utility of DERS-16 screenings and explore which PeriPD trajectories benefit most from ER-centered interventions. As a resilience factor, strengthened ER abilities can buffer stress, improve maternal mental health and ultimately support positive parent-child relationships and child development.^18,22,23,31^

## Acknowledgements

We thank Richard Aubrey White for the guidance in the statistical and visualization aspects of the study and Andreas Frick for discussions on the use of instruments to assess emotional regulation.

## Author Contribution

FW: Conceptualization; Methodology; Formal analysis; Investigation; Visualization; Writing – original draft. EF: Conceptualization; Validation; Writing – review & editing. BD: Conceptualization; Methodology; Funding acquisition; Supervision; Validation; Writing – review & editing. AS: Project administration; Conceptualization; Data acquisition; Methodology; Investigation; Funding acquisition; Resources; Supervision; Validation; Writing – review & editing.

## Data Availability Statement

Datasets used in the present study are available upon reasonable request from the corresponding authors.

