## Supplementary Information for "Emotion regulation is robustly associated with depressive symptoms across the peripartum – data from a prospective Swedish national cohort study"

### Results

#### Emotion regulation across the peripartum

Pearson correlation analysis showed that the DERS-16 total score assessed between 16-25 weeks antepartum was strongly correlated with DERS-16 total score assessed between 17-25 weeks postpartum ($r(197) = .655$, $p < .001$; see *Supplementary Figure 1).* Further, we conducted paired samples t-test to assess differences in DERS-16 total score between the peripartum assessment periods. While DERS-16 total score was slightly higher in the postpartum ($Mean_{postpartum} = 32.45$, $SD_{postpartum} = 13.67$) compared to the antepartum assessment ($Mean_{antepartum} = 31.08$, $SD_{antepartum} = 12.44$), no significant difference in DERS-16 total score between the two timepoints was found ($t(198) = -1.78$, $p = .076$).

#### Differences in sample characteristics between peripartum depression trajectories

Group differences in sample characteristics between the peripartum depression trajectories were seen for *age* ($F(4,129) = 2.62$, $p = .038$, $\eta p^2 = .08$), with the early postpartum onset group being of marginally higher age than the late postpartum onset group ($p = .056$). All other trajectory groups did not differ in age at registration (all *p*s > .231). Furthermore, differences were found for *self-reported depression history* (Fisher's Exact Test, $p = .009$), with a higher distribution of depression history in the early postpartum onset followed by the persistent depression group. Also, groups differed on *SOC-13 total score* ($F(4,128) = 4.13$, $p = .004$, $\eta p^2 = .11$), with the persistent depression group having lower scores than the healthy group ($p = .003$); *RS-14* ($F(4,126) = 22.11$, $p < .001$, $\eta p^2 = .41$), with the late postpartum onset having lower scores than the healthy group ($p = .003$), and the persistent having lower scores than the healthy, antepartum only, and early postpartum onset depression groups (all *p*s < .001); *VPSQ total score* ($H = 31.61$, $p < .001$), with higher scores in the early and late postpartum onset than the healthy group (all *p*s ≤ .016), and higher scores in the persistent than the healthy, antepartum only, and early postpartum onset groups (all *p*s ≤ .003); *FOBS* ($H = 12.48$, $p = .014$), with higher scores for the early and late postpartum onset (all *p*s ≤ .05) as well as higher scores for the persistent depression vs the healthy group ($p = .013$). No other differences between the groups were found for the psychometric scales (all *p*s ≥. 053). *Mode of delivery* differed between the trajectory groups (Fisher's Exact Test, $p$ = .008) with the healthy group having more vaginal deliveries, the antepartum only and late postpartum onset group having more planned caesarean sections, and the early and late postpartum as well as the persistent having more acute caesarean sections. Lastly, *delivery experience* differed between groups ($H$ = 12.65, $p$ = .013) with the healthy reporting better delivery experience vs the late postpartum onset and persistent depression groups (all *p*s ≤ .035) as well as the early postpartum onset reporting better delivery vs the persistent depression groups ($p$ = .048). No further differences were found between the trajectory groups for delivery experience (all *p*s > .098).

**Supplementary Table 1.** Linear regression models on Edinburgh Postnatal Depression Scale (EPDS) total score across the peripartum period (outcome).

| **(Outcome)** EPDS total score across the peripartum period | **Exposure** / Potential confounders | **(1)** **Unadjusted** | **(2)** **Adjusted for potential confounders – EPDS total score 12-22 w antepartum** | **(3)** **Adjusted for potential confounders + EPDS total score 12-22 w antepartum** |
|---|---|---|---|---|
| 24-34 w antepartum | | | $R^2$ = .42 *** | $R^2$ = .42 *** |
| | | | N = 423 | N = 423 |
| | **DERS-16** | .22 (.19 to .25) *** | .15 (.11 to .19) *** | .15 (.11 to .19) *** |
| | age | -.10 (-.19 to 0.00) *ns* | -.07 (-.17 to ,03) *ns* | -.07 (-.17 to ,03) *ns* |
| | BMI | .15 (.06 to .23) *** | .11 (.04 to .18) * | .11 (.04 to .18) * |
| | university education | -.1.06 (-2.01 to -.10) * | -0.05 (-.96 to .87) *ns* | -0.05 (-.96 to .87) *ns* |
| | parity | .24 (.01 to .46) * | .27 (.03 to .51) * | .27 (.03 to .51) * |
| | pregnancy loss | .78 (-.20 to 1.71) *ns* | .37 (-.58 to 1.31) *ns* | .37 (-.58 to 1.31) *ns* |
| | depression history | .76 (-.20 to 1.71) *ns* | .32 (-.54 to 1.19) *ns* | .32 (-.54 to 1.19) *ns* |
| | PMS | 3.10 (1.63 to 4.57) *** | 1.15 (-.20 to 2.50) *ns* | 1.15 (-.20 to 2.50) *ns* |
| | OC mood | .93 (.04 to 1.83) * | .41 (-.40 to 1.22) *ns* | .41 (-.40 to 1.22) *ns* |
| | FOBS 13-42 w ap | .07 (.05 to .08) *** | .04 (.02 to .06) *** | .04 (.02 to .06) *** |
| | RS-14 20-42 w ap | -.18 (-.21 to -.15) *** | -.06 (-.10 to -.01) * | -.06 (-.10 to -.01) * |
| | SOC-13 18-42 w ap | -.26 (-.32 to -.19) *** | -.06 (-.13 to .01) *ns* | -.06 (-.13 to .01) *ns* |
| | VPSQ 32-42 w ap | .37 (.25 to .48) *** | -.04 (.08 to .29) *ns* | -.04 (.08 to .29) *ns* |
| 36-42 w antepartum | | | $R^2$ = .43 *** | $R^2$ = .43 *** |
| | | | N = 365 | N = 365 |
| | **DERS-16** | .23 (.19 to .26) *** | .15 (.11 to .20) *** | .15 (.11 to .20) *** |
| | age | -.01 (-.13 to .11) *ns* | .03 (-.08 to .13) *ns* | .03 (-.08 to .13) *ns* |
| | BMI | .15 (.06 to .23) *** | .11 (.03 to .18) * | .11 (.03 to .18) * |
| | university education | -.60 (-1.75 to .56) *ns* | .06 (-.94 to 1.06) *ns* | .06 (-.94 to 1.06) *ns* |
| | parity | .19 (-.08 to .47) *ns* | .09 (-.17 to .36) *ns* | .09 (-.17 to .36) *ns* |
| | pregnancy loss | 1.31 (.16 to 2.45) * | .95 (-.09 to 1.99) *ns* | .95 (-.09 to 1.99) *ns* |
| | depression history | 3.16 (2.12 to 4.20) *** | 1.12 (.17 to 2.06) * | 1.12 (.17 to 2.06) * |
| | PMS | 2.64 (.86 to 4.42) * | .52 (-.95 to 2.00) *ns* | .52 (-.95 to 2.00) *ns* |
| | OC mood | 1.13 (.06 to 2.21) * | .61 (-.28 to 1.49) *ns* | .61 (-.28 to 1.49) *ns* |
| | FOBS 13-42 w ap | .07 (.05 to .09) *** | .04 (.02 to .06) *** | .04 (.02 to .06) *** |
| | RS-14 20-42 w ap | -.16 (-.20 to -.12) *** | 0.00 (-.05 to .04) *ns* | 0.00 (-.05 to .04) *ns* |
| | SOC-13 18-42 w ap | -.27 (-.35 to -.20) *** | -.09 (-.17 to -.02) * | -.09 (-.17 to -.02) * |
| | VPSQ 32-42 w ap | .47 (.35 to .60) *** | .10 (-.03 to .23) *ns* | .10 (-.03 to .23) *ns* |
| **BIRTH** | | | | |
| 1-4 w postpartum | | | $R^2$ = .33 *** | $R^2$ = .36 *** |
| | | | N = 332 | N = 312 |
| | **DERS-16** | .17 (.13 to .22) *** | .08 (.03 to .13) *** | .06 (.00 to .11) * |
| | age | -.01 (-.14 to 12) *ns* | .07 (-.05 to .20) *ns* | .09 (-.04 to .22) *ns* |
| | BMI | .09 (-.01 to .19) *ns* | .08 (-.01 to .17) *ns* | .06 (-.03 to .15) *ns* |
| | university education | .83 (-.42 to 2.09) *ns* | 1.0 (-.19 to 2.19) *ns* | .96 (-.26 to 2.18) *ns* |
| | parity | -.06 (-.36 to .24) *ns* | .01 (-.31 to .32) *ns* | -.04 (-.36 to .29) *ns* |

| | | | | |
|---|---|---|---|---|
| | pregnancy loss | .19 (-1.06 to 1.43) *ns* | .20 (-1.03 to 1.43) *ns* | .14 (-1.12 to 1.40) *ns* |
| | depression history | 1.96 (.80 to 3.12) *** | 0.00 (-1.13 to 1.13) *ns* | -.23 (-1.39 to .93) *ns* |
| | PMS | 3.80 (1.89 to 5.71) *** | 2.10 (.34 to 3.84) * | 2.19 (.40 to 3.98) * |
| | OC mood | 1.73 (.58 to 2.89) * | 1.32 (.27 to 2.37) * | 1.25 (.18 to 2.32) * |
| | FOBS 13-42 w ap | .04 (.02 to .06) *** | .01 (-.01 to .03) *ns* | .01 (-.01 to 0.3) *ns* |
| | RS-14 20-42 w ap | -.15 (-.20 to -.10) *** | -.03 (-.08 to .02) *ns* | -.01 (-.07 to .33) *ns* |
| | SOC-13 18-42 w ap | -.22 (-.30 to -.13) *** | -.03 (-.12 to .06) *ns* | -.02 (-.11 to .07) *ns* |
| | VPSQ 32-42 w ap | .55 (.41 to .68) *** | .28 (.13 to .43) *** | .26 (.11 to .42) *** |
| | delivery experience | -.08 (-.10 to -.05) *** | -.04 (-.07 to -.02) *** | -.05 (-.08 to -.02) *** |
| | neonatal issues 0-2 w pp | -.08 (-.10 to -.05) *** | 1.80 (.20 to 3.4) * | 1.92 (.28 to 3.56) * |
| | EPDS w 12-22 ap | .41 (-.30 to .52) *** | NA | .20 (.07 to .33) * |
| 6-13 w postpartum | | | $R^2$ = .34 *** | $R^2$ = .38 *** |
| | | | N = 282 | N = 262 |
| | **DERS-16** | .18 (.13 to .22) *** | .09 (.04 to .14) *** | .06 (.00 to .11) * |
| | age | -.10 (-.23 to .03) *ns* | -.04 (-.16 to .09) *ns* | -.01 (-.14 to .12) *ns* |
| | BMI | .11 (.01 to .21) * | .09 (0.00 to .18) * | -.06 (-.03 to .15) *ns* |
| | university education | .07 (-1.21 to 1.34) *ns* | .72 (-.48 to 1.93) *ns* | .67 (-.55 to 1.90) *ns* |
| | parity | -.09 (-.39 to .21) *ns* | .18 (-.14 to .49) *ns* | .13 (-.20 to .45) *ns* |
| | pregnancy loss | -1.25 (-2.51 to .01) *ns* | -1.50 (-2.75 to -.26) * | 1.58 (-2.85 to -.32) * |
| | depression history | 2.27 (1.10 to 3.44) *** | .71 (-.43 to 1.85) *ns* | .42 (-.75 to 1.58) *ns* |
| | PMS | 2.13 (.17 to 4.10) * | .03 (-1.75 to 1.80) *ns* | .16 (-1.65 to 1.96) *ns* |
| | OC mood | 1.67 (.49 to 2.85) * | 1.32 (.26 to 2.38) * | 1.23 (.15 to 2.31) * |
| | FOBS 13-42 w ap | .05 (.03 to .08) *** | .03 (.00 to .05) * | .02 (.00 to .04) * |
| | RS-14 20-42 w ap | -.16 (-.21 to -.12) *** | -.06 (-.12 to -.01) * | -.04 (-.09 to .02) *ns* |
| | SOC-13 18-42 w ap | -.21 (-.29 to -.12) *** | -.02 (-.11 to .07) *ns* | -.01 (-.10 to.09) *ns* |
| | VPSQ 32-42 w ap | .43 (.29 to .57) *** | .12 (-.03 to .27) *ns* | .10 (-.06 to .25) *ns* |
| | delivery experience | -.05 (-.08 to -.02) *** | -.02 (-.05 to .01) *ns* | -.03 (-.05 to 0.00) *ns* |
| | neonatal issues 0-2 w pp | -.05 (-.08 to -.02) *** | 1.15 (-.47 to 2.77) *ns* | 1.30 (-.35 to 3.0) *ns* |
| | EPDS w 12-22 ap | .46 (.36 to .57) *** | NA | .25 (.12 to .39) *** |
| 14-23 w postpartum | | | $R^2$ = .46 *** | $R^2$ = .57 *** |
| | | | N = 224 | N = 211 |
| | **DERS-16** | .21 (.17 to .26) *** | .15 (.09 to .20) *** | .10 (.04 to .15) *** |
| | age | -.05 (-.20 to 1.00) *ns* | .07 (-.06 to .21) *ns* | .12 (-.01 to .24) *ns* |
| | BMI | .18 (.07 to .29) * | .17 (-.07 to .26) *** | .13 (.04 to .21) * |
| | university education | -.65 (-2.12 to .82) *ns* | -.16 (-1.42 to 1.10) *ns* | -.24 (-1.42 to .94) *ns* |
| | parity | -.07 (-.42 to .27) *ns* | .08 (-.25 to .41) *ns* | -.01 (-.32 to .31) *ns* |
| | pregnancy loss | -.75 (-2.21 to .70) *ns* | -1.09 (-2.40 to .21) *ns* | -1.22 (-2.44 to .01) *ns* |
| | depression history | 1.72 (.36 to 3.09) * | -.12 (-1.31 to 1.07) *ns* | -.60 (-1.73 to .52) *ns* |
| | PMS | 1.27 (-1.01 to 3.55) *ns* | -.45 (-2.31 to 1.40) *ns* | -.24 (-1.98 to 1.50) *ns* |
| | OC mood | 1.90 (.55 to 3.26) * | 1.68 (.58 to 2.80) * | 1.54 (.50 to 2.58) * |
| | FOBS 13-42 w ap | .05 (.02 to .07) *** | .01 (-.01 to .03) *ns* | 0.00 (-.02 to .02) *ns* |
| | RS-14 20-42 w ap | -.19 (-.24 to -.14) *** | -.07 (-.13 to -.01) * | -.03 (-.09 to .03) *ns* |
| | SOC-13 18-42 w ap | -.21 (-.31 to -.11) *** | .01 (-.08 to .11) *ns* | .03 (-.06 to .12) *ns* |
| | VPSQ 32-42 w ap | .45 (.29 to .61) *** | .08 (-.08 to .24) *ns* | .04 (-.11 to .19) *ns* |
| | delivery experience | -.08 (-.12 to -.05) *** | -.06 (-.09 to -.03) *** | -.07 (-.10 to -.04) *** |
| | neonatal issues 0-2 w pp | -.08 (-.12 to -.05) *** | -2.00 (-3.67 to -.28) * | -1.72 (-3.31 to -.13) * |
| | EPDS w 12-22 ap | .60 (.49 to .71) *** | NA | 42 (.29 to .54) *** |

| | | | | |
|---|---|---|---|---|
| | | | $R^2$ = .34 *** | $R^2$ = .39 *** |
| | | | N = 155 | N = 149 |
| 24-35 w postpartum | **DERS-16** | .15 (.10 to .21) *** | .06 (-.01 to .13) + | .03 (-.05 to .10) *ns* |
| | age | -.05 (-.23 to .13) *ns* | .02 (-.16 to .20) *ns* | .05 (-.14 to .23) *ns* |
| | BMI | .15 (.02 to .29) * | .12 (0.00 to .25) *ns* | .10 (-.03 to .22) *ns* |
| | university education | -.17 (-1.93 to 1.59) *ns* | .40 (-1.33 to 2.12) *ns* | .34 (-1.38 to 2.07) *ns* |
| | parity | -.11 (-.53 to .30) *ns* | -.07 (-.52 to .39) *ns* | -.12 (-.58 to .33) *ns* |
| | pregnancy loss | -.08 (-1.82 to 1.67) *ns* | .01 (-1.78 to 1.79) *ns* | -.08 (-1.86 to 1.71) *ns* |
| | depression history | 2.50 (.89 to 4.11) * | 1.12 (-.52 to 2.75) *ns* | .80 (-.84 to 2.45) *ns* |
| | PMS | 2.12 (-.60 to 4.84) *ns* | .33 (-2.21 to 2.87) *ns* | .47 (-2.07 to 3.01) *ns* |
| | OC mood | 2.25 (.64 to 3.86) * | 2.12 (.60 to 3.64) * | 2.02 (.51 to 3.54) * |
| | FOBS 13-42 w ap | .06 (.03 to .09) *** | .03 (.00 to .06) * | .03 (-.01 to .06) *ns* |
| | RS-14 20-42 w ap | -.16 (-.23 to -.10) *** | -.08 (-.15 to 0.00) *ns* | -.05 (-.13 to .03) *ns* |
| | SOC-13 18-42 w ap | -.18 (-.30 to -.06) * | -.01 (-.13 to .12) *ns* | .01 (-.12 to .14) *ns* |
| | VPSQ 32-42 w ap | .40 (.20 to .59) *** | .07 (-.15 to .29) *ns* | .04 (-.18 to .26) *ns* |
| | delivery experience | -.07 (-.11 to -.03) * | -.04 (-.08 to -.01) * | -.05 (-.09 to -.01) * |
| | neonatal issues 0-2 w pp | -.07 (-.11 to -.03) * | -.52 (-2.85 to 1.80) *ns* | -.36 (-2.68 to 1.97) *ns* |
| | EPDS w 12-22 ap | .46 (.32 to .61) *** | NA | .27 (.09 to .46) * |
| | | | $R^2$ = .68 *** | $R^2$ = .76 *** |
| | | | N = 93 | N = 92 |
| 36-49 w postpartum | **DERS-16** | .22 (.15 to .29) *** | .09 (.02 to .16) * | .04 (-.02 to .11) *ns* |
| | age | -.13 (-.37 to .12) *ns* | -.05 (-.23 to .13) *ns* | -.02 (-.18 to .15) *ns* |
| | BMI | .23 (.06 to .41) * | .20 (.07 to .32) * | .16 (.05 to .27) * |
| | university education | -.91 (-3.32 to 1.49) *ns* | .02 (-1.68 to 1.71) *ns* | -.05 (-1.57 to 1.47) *ns* |
| | parity | -.08 (-.64 to .49) *ns* | .09 (-.36 to .54) *ns* | .02 (-.38 to .42) *ns* |
| | pregnancy loss | 0.00 (-2.38 to 2.39) *ns* | -.15 (-1.91 to 1.61) *ns* | -.25 (-.183 to 1.32) *ns* |
| | depression history | 3.44 (1.31 to 5.57) * | 1.71 (.10 to 3.32) * | 1.30 (-.15 to 2.75) *ns* |
| | PMS | 2.56 (-1.14 to 6.26) *ns* | .44 (-2.06 to 2.94) *ns* | .62 (-1.62 to 2.86) *ns* |
| | OC mood | 1.76 (-.46 to 3.98) *ns* | 1.46 (-.04 to 2.96) *ns* | 1.34 (-.01 to 2.68) *ns* |
| | FOBS 13-42 w ap | .08 (.04 to .12) *** | .05 (.02 to .08) * | .04 (.01 to .07) * |
| | RS-14 20-42 w ap | -.24 (-.31 to -.16) *** | -.09 (-.17 to -.01) * | -.06 (-.13 to .01) *ns* |
| | SOC-13 18-42 w ap | -.30 (-.45 to -.14) *** | -.06 (-.19 to .07) *ns* | -.05 (-.16 to .07) *ns* |
| | VPSQ 32-42 w ap | .53 (.28 to .78) *** | .06 (-.16 to .27) *ns* | .02 (-.17 to .22) *ns* |
| | delivery experience | -.10 (-.15 to -.04) *** | -.07 (-.11 to -.03) *** | -.08 (-.11 to -.04) *** |
| | neonatal issues 0-2 w pp | -.10 (-.15 to -.04) *** | -2.74 (-.5.03 to -.46) * | -2.53 (-4.58 to -.48) * |
| | EPDS w 12-22 ap | .65 (.48 to .81) *** | NA | .35 (.19 to .51) *** |

$^{ns}p \geq .196$, +$p = .111$, *$p < .050$, ***$p \leq .001$; ap: antepartum; pp: postpartum; w: weeks

*Note.* Models are corrected for (1) no other potential confounders variables (2) potential confounders excluding EPDS total score 12-22 weeks antepartum and (3) potential confounders including EPDS total score 12-22 weeks antepartum. $R^2$, N, beta coefficients, and 95% confidence interval are reported.

Included variables: DERS-16: Deficits in Emotion Regulation Scale-16 total score 16-25 weeks antepartum (exposure); BMI: pre-pregnancy BMI; university level education (vs less); parity; pregnancy loss: pregnancy loss history (vs never); depression history: self-reported depression history with professional help (vs no); PMS: past treatment for premenstrual disorder (vs never); OC mood: mental health issues due to oral contraceptives (vs never);

FOBS: Fear of Birth Scale, mean total score 13-42 weeks antepartum; RS-14: Resilience Scale-14 total score 20-42 weeks antepartum; SOC-13: Sense of coherence-13 total score 18-42 weeks antepartum; VPSQ: Vulnerable Personality Style Questionnaire total score 32-42 weeks antepartum (all potential confounders); Postpartum EPDS outcomes were additionally adjusted for delivery experience, neonatal issues up to two weeks postpartum, and in Model 3 also for EPDS: Edinburgh Postnatal Depression Scale total score at 12-22 weeks antepartum.

**Supplementary Table 2.** Edinburgh Postnatal Depression Scale (EPDS) mean total scores (standard deviation) for each assessment period across peripartum for the five distinct peripartum depression trajectory groups

| | **Peripartum depression trajectory group (based on EPDS scores across the peripartum period)** | | | | |
|---|---|---|---|---|---|
| | **Healthy** | **Antepartum only** | **Early postpartum onset** | **Late postpartum onset** | **Persistent** |
| **N** | 61 (45.4%) | 10 (7.5%) | 36 (26.9%) | 8 (6.0%) | 19 (14.2%) |
| **EPDS mean (sd) total score across the peri-partum period** | | | | | |
| 12-22 w ap | 5.0 (3.3) | 10.7 (3.9) | 6.2 (3.0) | 7.8 (2.8) | 14.9 (6.8) |
| 24-34 w ap | 4.4 (2.9) | 9.7 (5.4) | 6.1 (2.9) | 7.3 (2.8) | 14.2 (5.3) |
| 36-42 w ap | 3.6 (2.9) | 7.8 (4.0) | 7.5 (3.5) | 4.9 (3.5) | 15.6 (3.7) |
| **BIRTH** | | | | | |
| 1-4 w pp | 4.9 (3.1) | 5.6 (2.9) | 13.4 (4.5) | 6.7 (2.3) | 15.9 (4.9) |
| 6-13 w pp | 3.3 (2.9) | 3.9 (2.5) | 9.0 (4.5) | 6.0 (3.7) | 14.0 (4.3) |
| 14-23 w pp | 3.4 (3.2) | 6.1 (3.1) | 7.7 (4.3) | 11.6 (4.4) | 15.2 (4.2) |
| 24-35 w pp | 2.6 (2.4) | 3.6 (2.8) | 6.8 (5.5) | 11.5 (4.0) | 13.2 (4.0) |
| 36-42 w pp | 2.7 (2.8) | 3.7 (3.2) | 5.7 (3.5) | 11.0 (3.9) | 16.0 (1.8) |

**Supplementary Table 3.** Deficits of Emotion Regulation Scale-16 (DERS-16) mean (standard deviation) total score at 16-25 weeks antepartum and 17-25 weeks postpartum for the five distinct peripartum depression trajectory groups.

| | **Peripartum depression trajectory group (based on EPDS scores across the peripartum period)** | | | | |
|---|---|---|---|---|---|
| | **Healthy** | **Antepartum only** | **Early postpartum onset** | **Late postpartum onset** | **Persistent** |
| **N** | 61 (45.4%) | 10 (7.5%) | 36 (26.9%) | 8 (6.0%) | 19 (14.2%) |
| **DERS-16 mean (sd) total score** | | | | | |
| 16-25 w ap | 25.7 (7.9) | 31.0 (10.7) | 32.3 (10.8) | 37.5 (13.0) | 52.3 (12.5) |
| **BIRTH** | | | | | |
| 17-25 w pp | 25.9 (8.4) | 26.6 (5.2) | 30.4 (7.9) | 41.8 (16.4) | 49.2 (19.0) |
